# A single measurement of fecal hemoglobin concentration outperforms polygenic risk score in colorectal cancer risk assessment

**DOI:** 10.1101/2022.07.22.22277924

**Authors:** Tobias Niedermaier, Elizabeth Alwers, Xuechen Chen, Thomas Heisser, Michael Hoffmeister, Hermann Brenner

## Abstract

**PURPOSE:** Polygenic risk scores (PRS) have been proposed and are increasingly used for risk assessment in colorectal cancer (CRC) screening. Fecal immunochemical tests (FITs) are widely recommended and used as dichotomous tests for annual or biennial CRC screening, ignoring the quantitative information on fecal hemoglobin concentration above or below the positivity threshold.

**MATERIALS AND METHODS:** We aimed to assess and compare the ability of a single quantitative FIT and PRS to predict presence of advanced colorectal neoplasms (preclinical CRC or advanced adenoma) as a key indicator of CRC risk. A quantitative FIT (FOB Gold, Sentinel Diagnostics) was employed and a weighted PRS based on 140 common risk variants was determined among participants of screening colonoscopy in Germany. We compared areas under the curves (AUCs) of FIT and PRS for predicting presence of advanced colorectal neoplasm in the entire study population, and in subgroups defined by age, sex, family history of CRC, and history of colonoscopy.

**RESULTS:** A total of 3,025 participants aged 50-79 years were included, thereof 523 with advanced colorectal neoplasm and 2,502 participants without neoplasm. FIT clearly outperformed PRS in predicting presence of any advanced neoplasm in the entire study population (AUC 0.721, 95%CI 0.693-0.749 versus 0.591, 95%CI 0.564-0.617, p<0.0001), in younger (50-59 years) and older (60-79 years) participants, both sexes, those with and without a family history of CRC, and those with and without a previous colonoscopy (differences in AUC between 0.110 and 0.186, p=0.03 for those with previous colonoscopy and ≤0.005 for all other subgroups).

**CONCLUSION:** A single quantitative FIT, a low cost, easy-to-administer and universally available test, is more informative for CRC risk assessment than so far established PRS, irrespective of age, sex, family history, or history of colonoscopy.

## Introduction

Large-scale international genome-wide association studies have identified a rapidly increasing number of single nucleotide polymorphisms (SNPs) that are associated with colorectal cancer (CRC) risk (e.g.^1-4^). Even though associations of individual SNPs with CRC risk are mostly very small, integrative measures such as polygenic risk scores (PRS) based on the entity of the established SNPs have been shown to enable clinically relevant risk assessment that may help to develop novel, risk-adapted prevention and screening strategies. However, with areas under the curves (AUCs) of up to ∼0.60 to 0.65^4,5^, predictive performance has remained modest, and approaches with better risk discrimination are highly desirable in order to make risk prediction more accurate.

Most CRCs develop slowly over many years from advanced adenomas or other precancerous lesions, and biomarkers indicating presence of such precancerous lesions may be most promising candidates for CRC risk assessment. Fecal immunochemical tests (FITs), which quantify the concentration of hemoglobin in stool, are increasingly used for CRC screening^6^. Although their ability to detect precancerous lesions is lower than that for detecting CRC, AUCs for predicting presence of precancerous lesions in the order of 0.65-0.75 have been reported^7^, i.e., at higher levels than those reported for PRS in prediction of CRC risk. However, there is a lack of large-scale studies directly comparing performance of FIT and PRS in predicting presence of preclinical CRC or precancerous lesions. In this study, we aimed for such a head-to-head comparison in a large cohort of participants undergoing screening colonoscopy in Germany.

## Methods

This article follows the STARD (Standards for Reporting of Diagnostic Accuracy)^8^ and the FITTER (Faecal Immunochemical test for Haemoglobin Evaluation Reporting)^9^ guidelines.

### Study design and population

The analyses are based on data from the BliTz study, a large ongoing study on novel approaches to CRC screening among participants of screening colonoscopy which has been described in detail elsewhere^10-12^. In brief, participants of the German screening colonoscopy program are recruited in gastroenterology practices in Southern Germany. The program includes rigorous measures of quality control, and only highly experienced gastroenterologists are entitled to conduct screening colonoscopies. Prior to bowel preparation, stool and blood samples are taken for evaluating the ability of noninvasive tests to predict presence of CRC and its precursors using colonoscopy as reference standard. The study has been approved by the Ethics Committees of the Medical Faculty Heidelberg (178/2005) and of the responsible state physicians’ chambers [Baden-Württemberg, M118-05-f; Rheinland-Pfalz, 837.047.06(5145); Saarland, 217/13; Hessen, MC 254/2007] and is registered in the German Clinical Trials Register (DRKS-ID: DRKS00008737).

For the current analysis, we included participants recruited from November 2008 to January 2019. From November 2008 to February 2012, participants filled small containers with raw native fresh stool samples, stored them in provided plastic bags, kept them in the freezer and brought them to the practice visit at the time of colonoscopy. The stool-filled containers were immediately stored in the freezer (−15 to -40 °C) at the practice visit, shipped on dry ice to a central laboratory within one to few days, and analyzed with FOB Gold. From February 2012 to January 2019, participants collected stool samples in collection tubes containing hemoglobin stabilizing buffers (10 mg stool in 1.7 ml buffer, Sentinel Diagnostics, Milano, Italy; Ref. 11561H). The tubes were sealed in envelopes and mailed to the study center at the German Cancer Research Center, where they were kept at 2−8 °C in the refrigerator before transporting in a cold chain to the central laboratory (Labor Limbach, Heidelberg, Germany) for analysis with FOB Gold. It has been shown that diagnostic performance of FIT in the BliTz study was virtually identical between the 2 different sample handling methods used during the recruitment period ^13^ and that repeated thawing and freezing of stool samples had little impact on accuracy measures of FITs ^14^. One stool sample was collected per participant. Further details have been reported previously^15,16^.

### Genotyping and selection of single nucleotide polymorphisms (SNPs)

Genotyping was done in all participants in whom CRC or advanced adenomas were detected and a random sample of age- and sex-matched participants without advanced neoplasms. Blood cell DNA was genotyped using Illumina OncoArray-500k V1.0 BeadChip (Illumina, San Diego, CA, USA) for 305 subjects and Global Screening Array (Illumina, San Diego, CA, USA) for 3,730 of the finally included subjects. Genotyping quality was assessed as described previously^17^. Imputation of missing genotypes was performed uniformly using the Haplotype Reference Consortium (version r1.1.2016) as reference panel. PLINK v1.90 was used to extract SNPs for the required regions of interest.

### Construction of the polygenic risk score

For the PRS, we considered a recently reported set of 140 common risk variants that were associated with a higher risk of CRC in the world’s largest CRC genome-wide association study (GWAS) in populations of European descent^4^. A weighted PRS was constructed that accounted for both the numbers of risk alleles and their strength of association with CRC risk as reflected in their beta coefficients with CRC risk (see **Supplementary Table 1** for details). Blinding from colonoscopy results was ensured when the PRS was constructed and applied to the study participants.

### Laboratory analyses of FITs

Laboratory personnel were blinded with respect to colonoscopy findings. Abbott Architect c8000 was used for analyses of fecal samples by FOB Gold. The analytical range was 0.2-132 µg hemoglobin (Hb) per gram stool. The median time (inter-quartile range) between fecal sampling and laboratory analysis was 9 (6-13) days.

### Colonoscopy findings

Colonoscopy and histology reports were reviewed by trained investigators who were blinded with respect to genetic and stool testing results. Each participant was classified to one of the following categories according to the most advanced finding at colonoscopy: CRC, advanced adenoma, non-advanced adenoma, other or no finding. Adenomas were defined as advanced if they matched any of the following features: size ≥1 cm, tubulovillous or villous architecture, or high-grade dysplasia. Sessile serrated polyps were not considered as outcome because they were rarely diagnosed and reported as such during the earlier years of the recruitment period.

### Statistical analyses

Several exclusion criteria were applied in order to focus on average-risk screening participants and to minimize the potential of missed neoplasms at colonoscopy (age <50 or ≥80 years, history of CRC or inflammatory bowel disease, previous colonoscopy within the past 5 years, inadequate bowel preparation or incomplete colonoscopy). In addition, participants with only nonadvanced adenomas or undefined polyps, whose relevance with respect to CRC risk is less certain, were excluded from the main analyses in order to focus on the distinction of participants with advanced neoplasm and those without neoplasms. However, we carried out additional sensitivity analyses in which participants with only nonadvanced adenomas were kept in the study population and included in the group of participants not carrying advanced adenomas.

We first described the study population with respect to the distribution of sex, age, family history of CRC, previous colonoscopy, and most advanced finding at screening colonoscopy. Then, we assessed the ability of FIT and PRS, coded as continuous variables (fecal hemoglobin concentration and weighted numbers of risk alleles) to predict presence of advanced neoplasms (preclinical CRC or advanced adenoma). This outcome was selected as primary outcome in our analysis because patients with advanced neoplasm have by far the highest risk to be diagnosed with CRC in the following years in the absence of screening. We constructed receiver operating characteristics (ROC) curves to visualize the ability of FIT and PRS to distinguish those with advanced neoplasms from participants without neoplasms for the entire range of possible cutoffs. In addition to the main analysis in the entire study population, stratified ROC analyses were carried out among younger (50-59 years) and older (60-79 years) participants, men and women, those with and without a family history of CRC, and those with and without a previous colonoscopy. Discriminative ability was quantified by AUCs, which were compared between PRS and FIT by DeLong’s test^18^. Statistical analyses were conducted using R and the package “pROC”^19^. Tests for statistical significance were two-sided at an α level of 0.05.

## Results

### Study participants

The flow of study participants included in this analysis is shown in **Figure 1**. Of 5,368 participants with FIT and genotyping results, 1,333 were excluded to ensure representativeness of an average-risk screening population and to minimize the risk of missed neoplasms. In detail, exclusion criteria were age (<50 or ≥80 years, N=190), a history of CRC or inflammatory bowel disease (N=43), colonoscopy in the previous 5 years (N=359), inadequate bowel preparation (N=535), incomplete colonoscopy (N=39). In order to focus on the distinction of participants with advanced neoplasm and those without neoplasms, we also excluded participants with nonadvanced adenoma (N=1,010) or undefined polyps (N=167).

**Figure 1.**
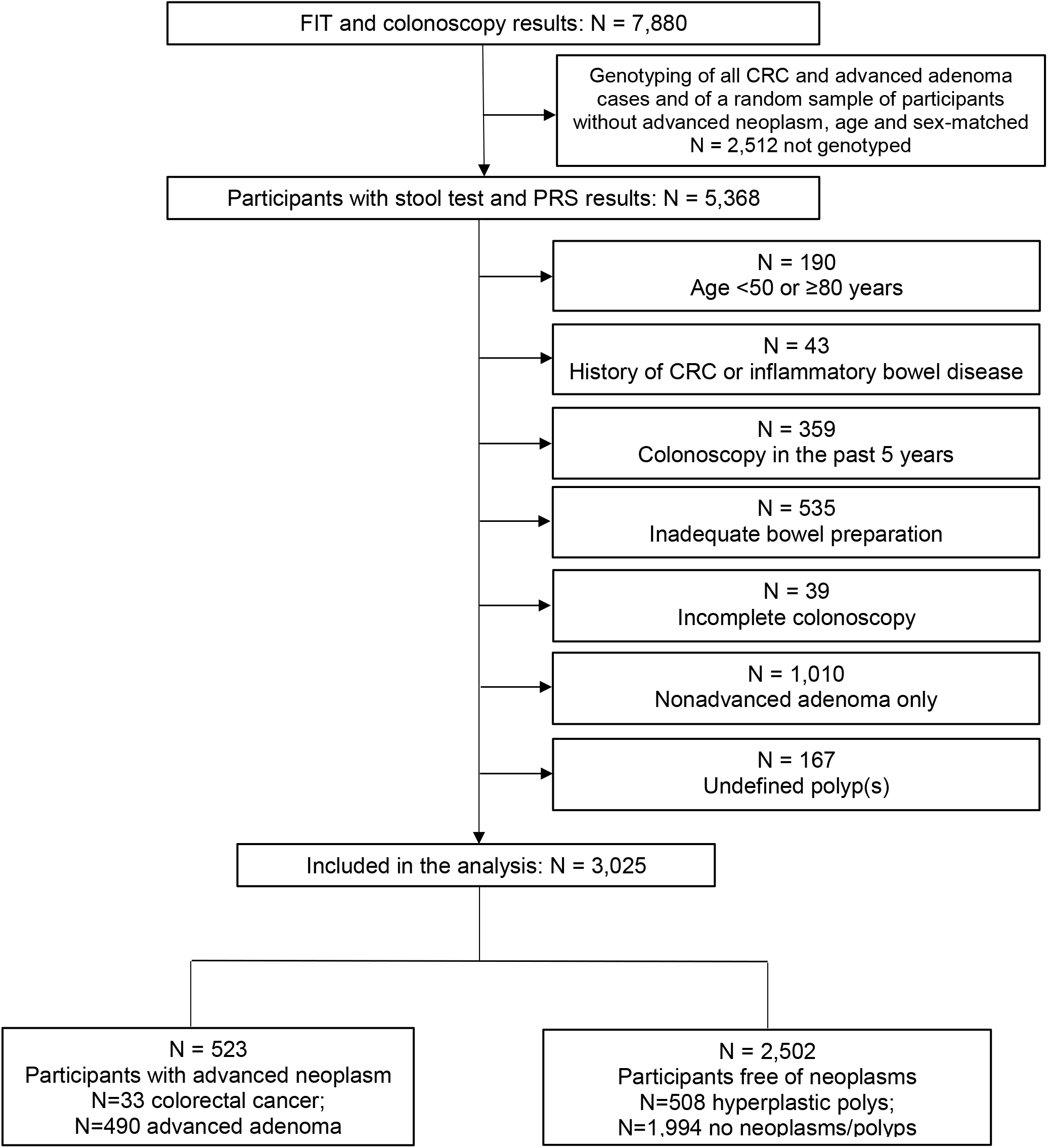
Flow chart of the study participants included in this analysis Abbreviations: FIT, fecal immunochemical test; CRC, colorectal cancer; PRS, polygenic risk score.

Characteristics of the study participants are summarized in **Table 1**. Among participants with advanced neoplasm (N=523), mean age and the proportion of males were 63.1 years and 61.0%. A family history of CRC was reported by 12.8% of participants with advanced neoplasm, and 22.0% of those 523 participants had had a previous colonoscopy. Among participants without neoplasm (N=2,502), mean age and the proportion of males were 61.3 years and 44.4%. A family history of CRC was reported by 12.2% of those 2,502 participants, and 33.5% reported having had a previous colonoscopy.

**Table 1.** Characteristics of the study population

| Characteristic |  | Any advanced neoplasm (N=523) | % | No neoplasm (N=2,502) | % |
| --- | --- | --- | --- | --- | --- |
| Sex | Male | 319 | 61.0 | 1,112 | 44.4 |
|  | Female | 204 | 39.0 | 1,390 | 55.6 |
| Age (years) | 50-54 | 17 | 3.3 | 121 | 4.8 |
|  | 55-59 | 193 | 36.9 | 1135 | 45.4 |
|  | 60-64 | 110 | 21.0 | 516 | 20.6 |
|  | 65-69 | 85 | 16.3 | 388 | 15.5 |
|  | 70-74 | 81 | 15.5 | 254 | 10.2 |
|  | 75-79 | 37 | 7.1 | 88 | 3.5 |
| Age (mean, standard deviation) |  | 63.1 (6.9) | .. | 61.3 (6.3) | .. |
| (Known) family history of colorectal cancer | Yes | 67 | 12.8 | 305 | 12.2 |
|  | No | 456 | 87.2 | 2,197 | 87.8 |
| Previous colonoscopy | Yes | 115 | 22.0 | 837 | 33.5 |
|  | No | 408 | 78.0 | 1,665 | 66.5 |

### ROC curves and AUCs

Figure 2 shows the ROC curves for predicting presence of any advanced neoplasm (preclinical colorectal cancer or advanced adenoma), compared to participants without neoplasms. FIT was clearly superior to PRS in predicting presence advanced neoplasms (AUC 0.721 vs. 0.591, p<0.0001). With inclusion of participants with only non-advanced adenomas in the group of participants without AN, AUCs of both FIT and PRS were slightly lower (0.710 and 0.574), whereas the difference was slightly larger (ΔAUC=0.136 vs. 0.130) and statistical significance remained unchanged (p<0.0001) (**Supplementary Figure 1**).

**Figure 2.**
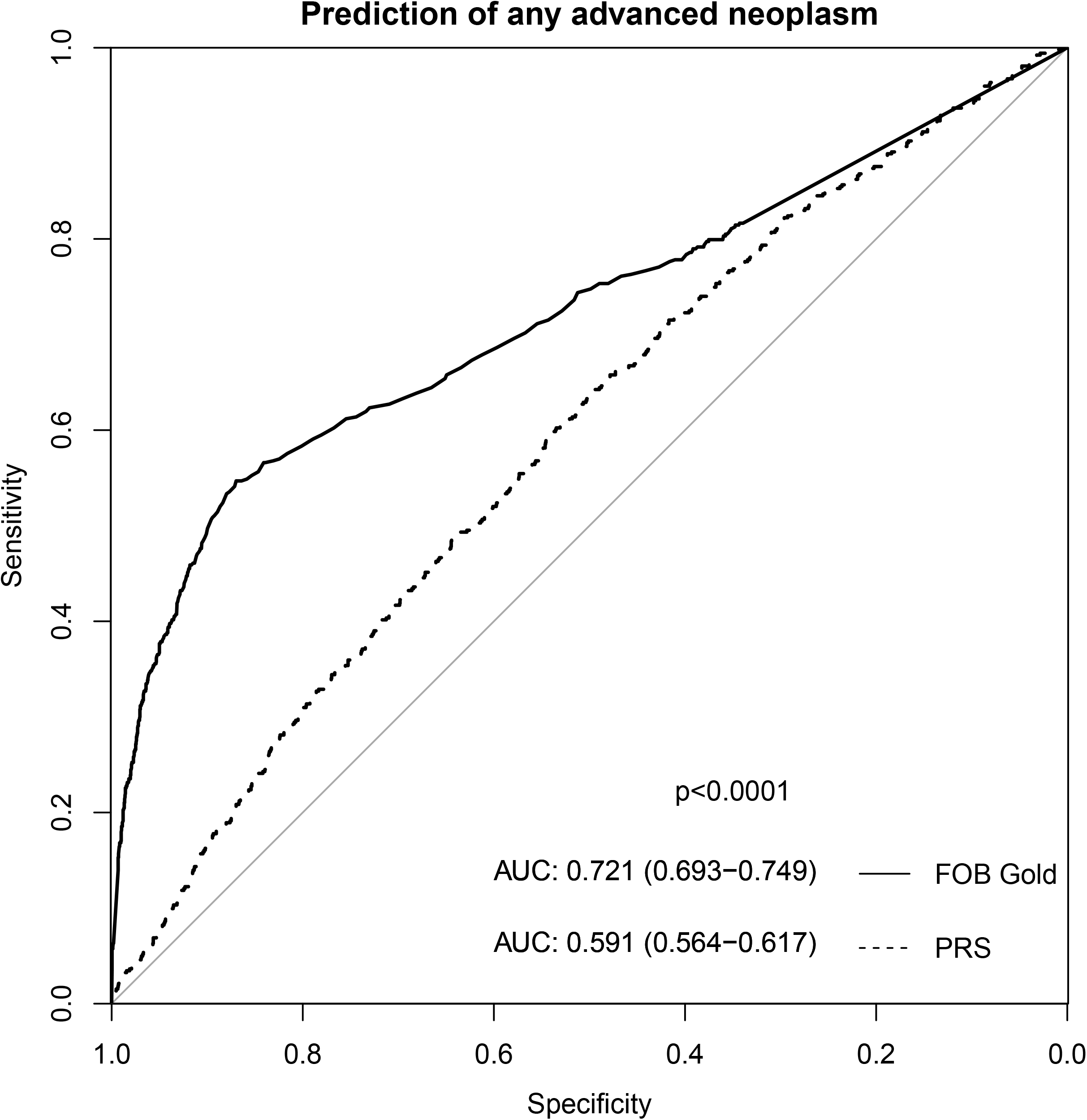
ROC curves, AUCs and p values for difference in AUCs between fecal immunochemical test (FOB Gold) and weighted polygenic risk score for prediction of any advanced colorectal neoplasm. Abbreviations: AUC, area under the receiver operating characteristics (ROC) curve; PRS, polygenic risk score; p, p value for difference in AUCs between fecal immunochemical test and PRS for predicting presence of any advanced colorectal neoplasm (DeLong’s test).

As shown in **Figures 3 and 4**, predictive performance of FIT clearly outperformed that of PRS in both younger (<60 years) and older (≥60 years) participants (ΔAUC=0.110 and 0.141, p=0.001 and <0.0001, respectively), men and women (ΔAUC=0.127 and 0.118, respectively, p<0.0001 and p=0.0003, respectively), those with and without a family history of CRC (ΔAUC=0.186 and 0.120, p=0.004 and p<0.0001, respectively), and those with and without a previous colonoscopy (ΔAUC=0.091 and 0.144, p=0.03 and p<0.0001, respectively).

**Figure 3.**
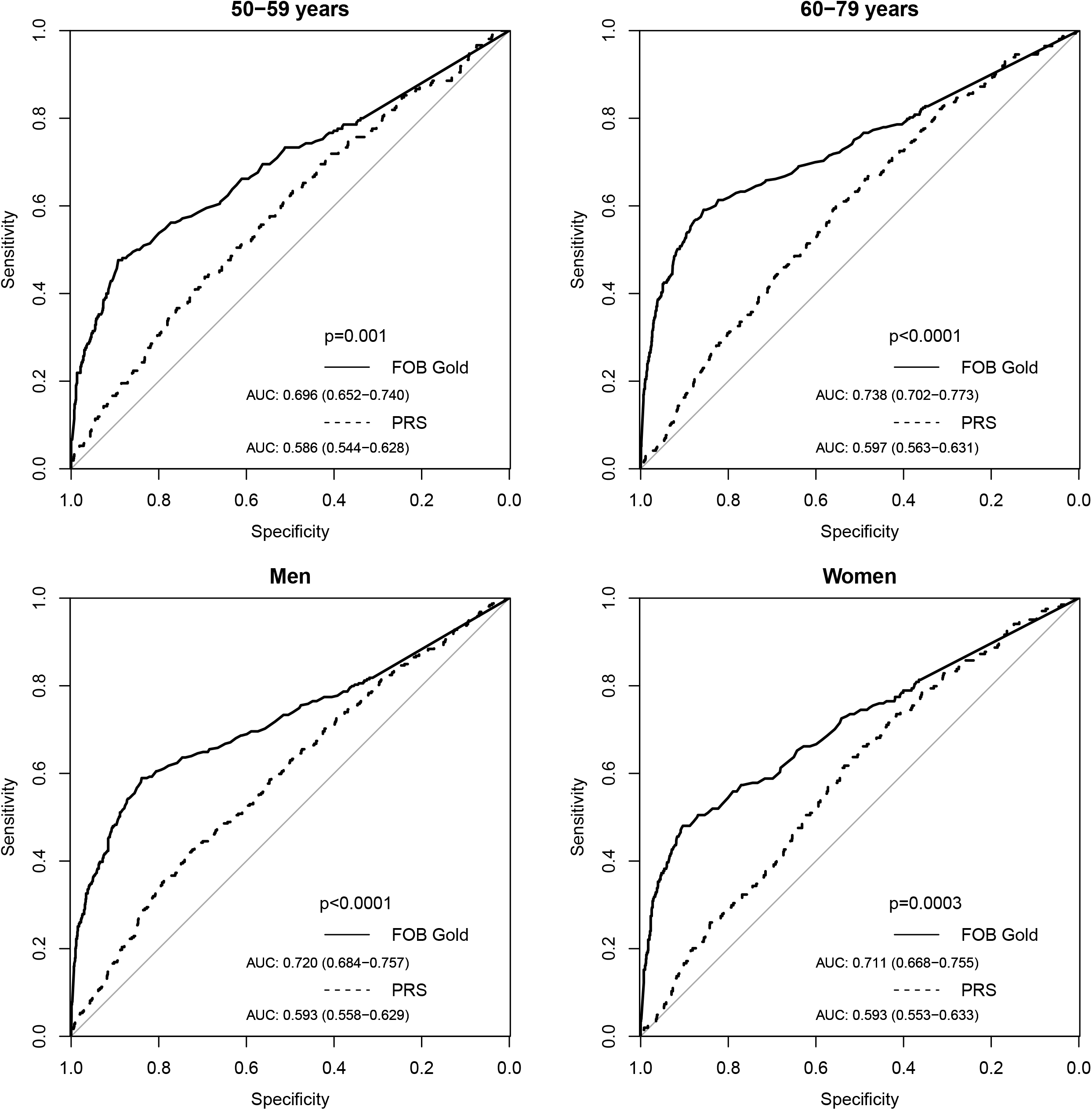
ROC curves, AUCs and p values for difference in AUCs between fecal immunochemical test (FOB Gold) and weighted polygenic risk score for predicting presence of advanced colorectal neoplasms, stratified by age and sex. Abbreviations: ROC, receiver operating characteristics; AUC, area under the ROC curve; PRS, polygenic risk score; p, p-value for difference in AUCs between fecal immunochemical test and PRS for predicting presence of any advanced colorectal neoplasm (DeLong’s test).

**Figure 4.**
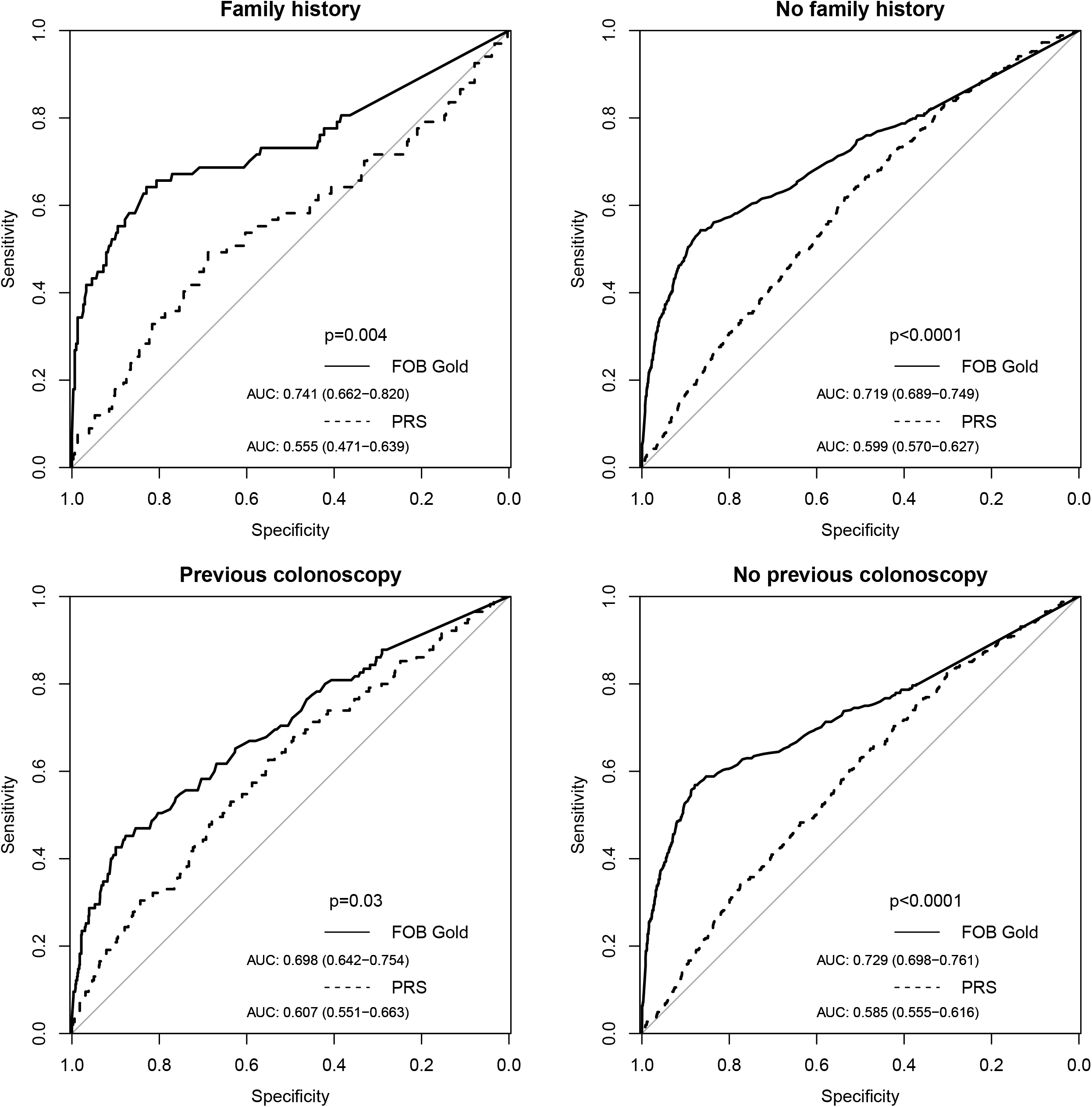
ROC curves, AUCs and p values for difference in AUCs between fecal immunochemical test (FOB Gold) and weighted polygenic risk score for predicting presence of advanced colorectal neoplasms, stratified by family history and history of previous colonoscopy. Abbreviations: ROC, receiver operating characteristics; AUC, area under the ROC curve; colo, colonoscopy; PRS, polygenic risk score; p, p-value for difference in AUCs between fecal immunochemical test and PRS for predicting presence of advanced colorectal neoplasms (DeLong’s test).

## Discussion

In this study among >3,000 participants of screening colonoscopy, we provide a head-to-head comparison of the ability of a quantitative FIT and a weighted PRS to predict presence of any advanced colorectal neoplasm, the best established proxy of CRC risk, in the target population for CRC screening. The FIT clearly outperformed the weighted PRS in predicting presence of advanced neoplasms, overall and in any of the assessed subgroups defined by age, sex, family history of CRC, and history of colonoscopy. These results suggest that a single quantitative measurement of fecal hemoglobin by quantitative FIT, a low cost, easy-to-administer and universally available test, can be a more informative tool for CRC risk assessment than genetic testing with PRS based on currently identified risk loci.

Our results on performance of either FIT or PRS for predicting presence of colorectal neoplasms are in line with previous studies^20-25^. However, our study strongly expands existing evidence by its large sample size, verification of presence or absence of neoplasms by colonoscopy among all participants in a true screening setting, and direct, head-to-head comparison of FIT and PRS. To limit complexity, we focused on use of either FIT or PRS as individual tools for risk assessment. Further improvement in risk prediction might be achieved for both approaches by combination with other information, such as risk factor based scores^26,27^, or other biomarkers or biomarker signatures which should be explored in further research.

Use of FIT is widely recommended and employed in an increasing number of countries for regular, i.e. annual or biennial, CRC screening in the average risk population. In this context, FITs are used as dichotomous screening tests, where a small proportion of participants with fecal hemoglobin concentration above a pre-defined positivity threshold are referred to colonoscopy, whereas the vast majority (typically ∼95%) of participants with lower fecal hemoglobin concentrations are invited to repeat screening after one or two years. Through exclusive use as a dichotomous test, most of the quantitative information on hemoglobin concentrations is simply discarded. Our results suggest that this is an enormous waste of information, as fecal hemoglobin concentrations seem to be highly informative on CRC risk not only for the small minority of participants above the positivity threshold, but also for “FIT = negative” participants, i.e., the vast majority of screening participants. Our results suggest that fecal hemoglobin concentration among the latter may still be very useful for risk assessment and might lend itself to more personalized, risk adapted screening strategies, much more so than the PRS that has received much more attention in this context. One possible application that has been suggested previously^28^ would be to define personalized screening intervals which might be reduced to one year in those with “high negative FIT results” and extended to more than two years among those with “low negative FIT results”.

Future studies might also investigate the potential for optimized FIT-based screening by considering quantitative information of two or more consecutive tests, such as cumulative hemoglobin concentrations. In a large longitudinal study, Grobbee et al.^29^ demonstrated that combined quantitative information of two negative FITs (sum of FIT concentrations) was a much stronger predictor of advanced neoplasia risk than age and sex. An approximately 12-fold higher risk of CRC, advanced adenoma, or any advanced neoplasm was found by Senore et al.^30^ in those with cumulative Hb concentrations of ≥20 µg/g compared to those with repeatedly 0 µg/g. Even better prediction might be achievable when assessing the change in Hb concentrations. Increases in stool hemoglobin concentration from 0 to a value somewhat below the cutoff within one year which could be an indicator for a more fast-growing lesion and suggest a higher risk of advanced neoplasia than FIT results that remain on a medium level for many years^31^. Such potential for “longitudinal” risk assessment is another advantage of FIT over PRS and should be investigated in more detail in future studies.

Another possible application for risk assessment could be use of a single FIT at the starting age of CRC screening, such as 45 or 50 years, to provide personalized, risk adapted recommendations to enter either colonoscopy based screening (typically to be repeated in 10-year intervals) or regular FIT based screening in countries where both options are offered, such as the US, Germany, Austria, or the Czech Republic. In this context, a much lower positivity threshold could be considered than in regular, annual or biennial FIT based screening.

Yet another possible application that has likewise previously been suggested for PRS, might be the definition of a personalized starting age of regular screening. For example, rather than determining PRS at an age at which initiation of screening might be considered for those at higher risk, such as 35 or 40 years, in order to define a personalized starting age, one might consider offering a quantitative “risk assessment FIT” every five years, say at ages 35, 40, 45, 50, 55, and 60, and recommend transition to regular screening (e.g. by colonoscopy every 10 years or annual or biennial FIT) once a certain hemoglobin concentration is surpassed.

Apart from superior performance for risk assessment, FIT has a number of further advantages over PRS when used in practice: Costs of conducting a FIT are substantially lower than that of genotyping. For example, in the US a typical FIT costs ∼$24-40^32,33^, whereas determination of a PRS was offered for ∼$200 in 2019^34^, and FITs are offered at even much lower costs in many other countries. Another clear advantage of (quantitative) fecal immunochemical testing is that it can be done in virtually all standard laboratories. Also, genotyping data are much more sensitive than FIT data, and require consulting of screenees by highly qualified medical personnel in how test results are to be interpreted. For example, in Germany, personal consultation by a specialist in human genetics is required by law for any genetic testing in medical practice. Finally, the sensitive nature of genetic data also requires more strict measures of data protection to prevent any kind of discrimination, e.g. with respect to insurance policies or at work. In particular, adherence to data safety regulations may be questionable or at least hard to prove for private companies that increasingly offer genotyping and grant access to results over the internet. An advantage of PRS-based over FIT-based risk assessment could be that genotyping by a genome-wide scan could enable simultaneous risk assessment for a variety of diseases (including but not restricted to various cancers) whereas FIT-based risk assessment is restricted to CRC risk.

Major strengths of our study were the large sample size which enabled estimation of AUCs with high precision, and conducting the study in a true screening setting, with verification of presence or absence of advanced neoplasms by screening colonoscopy in all participants. The large sample size enabled differentiated analyses of risk assessment by subgroups for which such risk assessment might be particularly relevant.

Our study also has limitations. Participants aged <50 years, for whom polygenic risk assessment might be particularly interesting, were not included because CRC screening is offered and typically done from age 50 onward in Germany. Results of this study are based on one particular FIT (FOB Gold) and one PRS and might differ slightly with other FITs and PRSs. In general, however, predictive performance of various FITs has been demonstrated to be very similar across different FIT manufacturers ^11^, and the PRS was derived from the world’s largest CRC GWAS in populations of European descent ^4^. Despite being considered as the gold standard for detecting colorectal neoplasms, screening colonoscopy is not perfect and likely to have missed some proportion of small, non-advanced neoplasms. However, we focused on advanced neoplasms as outcome which are rarely missed by colonoscopy^35,36^. Our study was restricted to a population of European ancestry which limits generalizability to other populations. Finally, our study was cross-sectional and risk of developing CRC could not be determined directly in a longitudinal manner. However, AUCs of PRS for predicting CRC have been consistently found to be in the order of 0.60 to 0.65 in large-scale cohort and case-control studies with CRC endpoints^4,5^, i.e., far below the AUC of 0.72 found for any advanced neoplasms in our study.

Despite its limitations, our study demonstrates that even a single FIT by far outperforms PRS in risk assessment for the presence of advanced colorectal neoplasms, a strong indicator for those at highest risk of developing CRC in the following years and most likely benefitting from CRC screening. Use of a single quantitative FIT, paying attention to hemoglobin concentrations also below the high positivity thresholds commonly employed in annual or biennial FIT-based screening, might be a more informative, economic, and feasible approach to risk assessment in CRC screening than genetic testing with currently available PRS. Further research should aim for exploring possibilities of further enhancing CRC risk assessment, e.g. by “longitudinal risk assessment” with FIT, combining FIT with risk factor information or other predictive biomarkers, and for evaluating the best possible use of risk assessment for effective and cost-effective CRC screening. Latter should be addressed in comprehensive modeling approaches, for which our study may provide important input parameters.

## Data Availability

Data availability: All data produced in the present study are available upon reasonable request to the authors.

## Abbreviations

AUC: area under the curve
FIT: fecal immunochemical test
PRS: polygenic risk score
ROC: receiver operating characteristics
CRC: colorectal cancer
GWAS: genome-wide association study
SNP: single nucleotide polymorphism.

