## Supplement for "A single measurement of fecal hemoglobin concentration outperforms polygenic risk score in colorectal cancer risk assessment"

**Supplementary Table 1.** Single-nucleotide polymorphisms considered to generate the polygenic risk score (PRS), adapted from Thomas et al. ^1^.

| **Locus** | **rsID lead variant** | **Variant** | **Chr.** | **Position (Build 37)** | **Risk allele** | **Reported beta coefficient risk allele** |
| --- | --- | --- | --- | --- | --- | --- |
| 1p34.3 | rs4360494 | 1:38455891_G/C | 1 | 38455891 | G | 0.0379 |
| 1p32.3 | rs12144319 | 1:55246035_T/C | 1 | 55246035 | C | 0.0661 |
| 1p36.12 | rs72647484 | 1:22587728_T/C | 1 | 22587728 | T | 0.0504 |
| 1p31.3 | rs7542665 | 1:62673037_T/C | 1 | 62673037 | C | 0.0334 |
| 1q25.3 | rs6678517 | 1:183002639_A/G | 1 | 183002639 | A | 0.073 |
| 1q41 | rs17011141 | 1:222112634_A/G | 1 | 222112634 | G | 0.0877 |
| 2q24.2 | rs448513 | 2:159964552_T/C | 2 | 159964552 | C | 0.0054 |
| 2q33.1 | rs11884596 | 2:199612407_T/C | 2 | 199612407 | C | 0.0342 |
| 2q33.1 | rs983402 | 2:199781586_T/C | 2 | 199781586 | T | 0.0622 |
| 2p16.3 | rs7606562 | 2:48686695_T/A | 2 | 48686695 | T | 0.0414 |
| 2q11.2 | rs11692435 | 2:98275354_G/A | 2 | 98275354 | G | 0.0492 |
| 2q35 | rs3731861 | 2:219191256_T/C | 2 | 219191256 | T | 0.0613 |
| 3q22.2 | rs10049390 | 3:133701119_G/A | 3 | 133701119 | A | 0.0455 |
| 3q13.2 | rs13086367 | 3:112903888_A/G | 3 | 112903888 | A | 0.0463 |
| 3q13.2 | rs72942485 | 3:112999560_G/A | 3 | 112999560 | G | 0.0545 |
| 3p21.1 | rs9831861 | 3:53088285_T/G | 3 | 53088285 | G | 0.0294 |
| 3p22.1 | rs35470271 | 3:40915239_A/G | 3 | 40915239 | G | 0.0994 |
| 3q13.2 | rs12635946 | 3:112916918_C/T | 3 | 112916918 | C | 0.0334 |
| 3q22.2 | rs113569514 | 3:133748789_T/C | 3 | 133748789 | T | 0.0414 |
| 3q26.2 | rs9876206 | 3:169517436_C/T | 3 | 169517436 | C | 0.0453 |
| 3p14.1 | rs6781752 | 3:66365163_G/A | 3 | 66365163 | A | 0.0597 |
| 4q31.21 | rs11727676 | 4:145659064_T/C | 4 | 145659064 | C | 0.0093 |
| 4q24 | rs1391441 | 4:106128760_G/A | 4 | 106128760 | A | 0.0148 |
| 4q22.2 | rs13149359 | 4:94938618_C/A | 4 | 94938618 | A | 0.052 |
| 5p13.1 | rs7708610 | 5:40102443_G/A | 5 | 40102443 | A | 0.0384 |
| 5p15.33 | rs78368589 | 5:1240204_C/T | 5 | 1240204 | T | 0.0786 |
| 5q21.1 | rs145364999 | 5:98206082_T/A | 5 | 98206082 | T | 0.3496 |
| 5p15.33 | rs2735940 | 5:1296486_A/G | 5 | 1296486 | G | 0.0865 |
| 5p13.1 | rs12514517 | 5:40280076_G/A | 5 | 40280076 | A | 0.1013 |

**Supplementary Table 1**, continued.

| **Locus** | **rsID lead variant** | **Variant** | **Chr.** | **Position (Build 37)** | **Risk allele** | **Reported beta coefficient risk allele** |
| --- | --- | --- | --- | --- | --- | --- |
| 5q22.2 | rs755229494 | 5:112097351_A/G | 5 | 112097351 | G | 0.6286 |
| 5q23.2 | rs12659017 | 5:125988175_G/A | 5 | 125988175 | G | 0.0374 |
| 5q31.1 | rs4976270 | 5:134467220_C/T | 5 | 134467220 | C | 0.0693 |
| 6p12.1 | rs13204733 | 6:55566108_A/G | 6 | 55566108 | G | 0.0643 |
| 6p21.33 | rs116685461 | 6:31315512_G/A | 6 | 31315512 | G | 0.0655 |
| 6p21.32 | rs9271695 | 6:32593080_A/G | 6 | 32593080 | G | 0.0889 |
| 6p21.33 | rs2516420 | 6:31449620_C/T | 6 | 31449620 | C | 0.1091 |
| 6p21.33 | rs116353863 | 6:31010185_T/C | 6 | 31010185 | C | 0.1202 |
| 6p21.31 | rs16878812 | 6:35569562_A/G | 6 | 35569562 | A | 0.0778 |
| 6p21.2 | rs9470361 | 6:36623379_G/A | 6 | 36623379 | A | 0.054 |
| 6p12.1 | rs62404966 | 6:55712124_C/T | 6 | 55712124 | C | 0.0724 |
| 6p21.33 | rs3131043 | 6:30758466_A/G | 6 | 30758466 | G | 0.0294 |
| 6p24.1 | rs2070699 | 6:12292772_G/T | 6 | 12292772 | T | 0.0294 |
| 6p22.1 | rs1476570 | 6:29809860_G/A | 6 | 29809860 | A | 0.0492 |
| 6p21.32 | rs3830041 | 6:32191339_C/T | 6 | 32191339 | T | 0.0645 |
| **6q21** | **rs6928864^a^** | **6:105966894_C/A** | **6** | **105966894** | **C** | 0.0531 |
| 6p21.1 | rs62396735 | 6:41702582_C/T | 6 | 41702582 | C | 0.033 |
| 7p13 | rs12672022 | 7:45136423_T/C | 7 | 45136423 | T | 0.0067 |
| 7p12.3 | rs80077929 | 7:46094089_C/T | 7 | 46094089 | T | 0.0093 |
| 7p12.3 | rs10951878 | 7:46926695_C/T | 7 | 46926695 | C | 0.0531 |
| 7p12.3 | rs3801081 | 7:47511161_A/G | 7 | 47511161 | G | 0.0253 |
| 8q24.21 | rs7013278 | 8:128414892_T/C | 8 | 128414892 | T | 0.0091 |
| 8q24.21 | rs4313119 | 8:128571855_G/T | 8 | 128571855 | G | 0.0518 |
| 8q23.3 | rs16892766 | 8:117630683_A/C | 8 | 117630683 | C | 0.2099 |
| 8q23.3 | rs6469654 | 8:117632965_G/C | 8 | 117632965 | G | 0.0677 |
| 8q24.11 | rs117079142 | 8:117790914_C/A | 8 | 117790914 | A | 0.1139 |
| 8q24.21 | rs6983267 | 8:128413305_G/T | 8 | 128413305 | G | 0.1052 |
| 9q22.33 | rs34405347 | 9:101679752_T/G | 9 | 101679752 | T | 0.0089 |
| 9p21.3 | rs1537372 | 9:22103183_G/T | 9 | 22103183 | G | 0.012 |
| 9q31.3 | rs10980628 | 9:113671403_T/C | 9 | 113671403 | C | 0.0511 |
| 10p14 | rs12217641 | 10:8663875_C/T | 10 | 8663875 | C | 0.0069 |
| 10q24.2 | rs10786560 | 10:101315166_G/A | 10 | 101315166 | G | 0.0082 |
| 10q22.3 | rs1250567 | 10:81046265_T/C | 10 | 81046265 | C | 0.047 |
| 10p14 | rs11255841 | 10:8739580_T/A | 10 | 8739580 | T | 0.1064 |
| 10q11.23 | rs10821907 | 10:52648454_C/T | 10 | 52648454 | C | 0.073 |
| 10q22.3 | rs704017 | 10:80819132_A/G | 10 | 80819132 | G | 0.0765 |
| 10q24.2 | rs11190164 | 10:101351704_A/G | 10 | 101351704 | G | 0.0889 |
| 10q25.2 | rs12246635 | 10:114288619_T/C | 10 | 114288619 | C | 0.0975 |
| 10q25.2 | rs11196170 | 10:114722621_G/A | 10 | 114722621 | A | 0.0527 |
| 11q13.4 | rs7946853 | 11:74409077_T/C | 11 | 74409077 | C | 0.0119 |

**Supplementary Table 1**, continued.

| **Locus** | **rsID lead variant** | **Variant** | **Chr.** | **Position (Build 37)** | **Risk allele** | **Reported beta coefficient risk allele** |
| --- | --- | --- | --- | --- | --- | --- |
| 11q22.1 | rs55864876 | 11:100717136_G/A | 11 | 100717136 | G | 0.015 |
| 11q22.1 | rs2186607 | 11:101656397_T/A | 11 | 101656397 | T | 0.0483 |
| 11q13.4 | rs61389091 | 11:74427921_C/T | 11 | 74427921 | C | 0.1934 |
| 1p15.4 | rs4450168 | 11:10286755_A/C | 11 | 10286755 | C | 0.0413 |
| 11q12.2 | rs174533 | 11:61549025_G/A | 11 | 61549025 | G | 0.0636 |
| 11q13.4 | rs7121958 | 11:74280012_T/G | 11 | 74280012 | G | 0.078 |
| 11q23.1 | rs3087967 | 11:111156836_T/C | 11 | 111156836 | T | 0.1122 |
| 12q13.3 | rs4759277 | 12:57533690_C/A | 12 | 57533690 | A | 0.0285 |
| 12q24.21 | rs1427760 | 12:115100714_T/C | 12 | 115100714 | C | 0.0424 |
| 12p13.32 | rs3217874 | 12:4400808_C/T | 12 | 4400808 | T | 0.0453 |
| 12p13.31 | rs10849433 | 12:6406904_T/C | 12 | 6406904 | C | 0.0468 |
| 12q12 | rs11610543 | 12:43134191_A/G | 12 | 43134191 | G | 0.0474 |
| 12p13.32 | rs35808169 | 12:4368607_T/C | 12 | 4368607 | C | 0.089 |
| 12p13.32 | rs3217810 | 12:4388271_C/T | 12 | 4388271 | T | 0.1181 |
| 12p13.31 | rs2250430 | 12:6421174_A/T | 12 | 6421174 | T | 0.0597 |
| 12p11.21 | rs77969132 | 12:31594813_C/T | 12 | 31594813 | T | 0.1583 |
| 12q13.12 | rs12372718 | 12:51171090_A/G | 12 | 51171090 | G | 0.0896 |
| 12q24.12 | rs597808 | 12:111973358_A/G | 12 | 111973358 | G | 0.0737 |
| 12q24.21 | rs7300312 | 12:115890922_T/C | 12 | 115890922 | C | 0.066 |
| 12p13.2 | rs2710310 | 12:12035649_C/T | 12 | 12035649 | C | 0.0145 |
| 13q22.1 | rs78341008 | 13:73791554_T/C | 13 | 73791554 | C | 0.0109 |
| 13q34 | rs8000189 | 13:111075881_C/T | 13 | 111075881 | T | 0.0473 |
| 13q22.1 | rs45597035 | 13:73649152_A/G | 13 | 73649152 | A | 0.0495 |
| 13q22.1 | rs1924816 | 13:73997961_A/G | 13 | 73997961 | A | 0.0506 |
| 13q13.3 | rs7333607 | 13:37462010_A/G | 13 | 37462010 | G | 0.0758 |
| 13q22.3 | rs1330889 | 13:78609615_T/C | 13 | 78609615 | C | 0.0453 |
| 13q13.2 | rs377429877 | 13:34092164_C/T | 13 | 34092164 | C | 0.0468 |
| 14q22.2 | rs1951864 | 14:54369299_G/A | 14 | 54369299 | A | 0.0059 |
| 14q23.1 | rs17094983 | 14:59189361_G/A | 14 | 59189361 | G | 0.0062 |
| 14q23.1 | rs8020436 | 14:59208437_G/A | 14 | 59208437 | A | 0.0294 |
| 14q22.2 | rs35107139 | 14:54419106_A/C | 14 | 54419106 | C | 0.0912 |
| 14q22.2 | rs4901473 | 14:54445157_G/A | 14 | 54445157 | G | 0.0465 |
| 15q23 | rs745213 | 15:68060389_T/G | 15 | 68060389 | G | 0.0072 |
| 15q22.31 | rs12594720 | 15:67007018_C/G | 15 | 67007018 | C | 0.0246 |
| 15q22.33 | rs56324967 | 15:67402824_T/C | 15 | 67402824 | C | 0.0689 |
| 15q13.3 | rs17816465 | 15:33156386_G/A | 15 | 33156386 | A | 0.069 |
| 15q13.3 | rs12708491 | 15:32992836_G/A | 15 | 32992836 | G | 0.0464 |
| 15q13.3 | rs2293581 | 15:33010736_G/A | 15 | 33010736 | A | 0.1248 |
| 15q26.1 | rs7495132 | 15:91172901_C/T | 15 | 91172901 | T | 0.0453 |
| 16q23.2 | rs9930005 | 16:80043258_C/A | 16 | 80043258 | C | 0.0061 |

**Supplementary Table 1**, continued.

| **Locus** | **rsID lead variant** | **Variant** | **Chr.** | **Position (Build 37)** | **Risk allele** | **Reported beta coefficient risk allele** |
| --- | --- | --- | --- | --- | --- | --- |
| 16q24.1 | rs12447408 | 16:86252544_G/A | 16 | 86252544 | A | 0.0079 |
| 16q22.1 | rs9924886 | 16:68743939_A/C | 16 | 68743939 | A | 0.055 |
| 16q24.1 | rs12149163 | 16:86339315_T/C | 16 | 86339315 | T | 0.0487 |
| 16q24.1 | rs62042090 | 16:86703949_C/T | 16 | 86703949 | T | 0.0481 |
| 17q24.3 | rs983318 | 17:70413253_G/A | 17 | 70413253 | A | 0.0397 |
| 17p13.3 | rs73975586 | 17:814243_A/T | 17 | 814243 | A | 0.0497 |
| 17p12 | rs1078643 | 17:10707241_G/A | 17 | 10707241 | A | 0.0747 |
| 17q25.3 | rs75954926 | 17:81061048_A/G | 17 | 81061048 | G | 0.0882 |
| 17q25.3 | rs373585858 | 17:80394556_G/A | 17 | 80394556 | A | 0.1103 |
| 17p13.3 | rs4968127 | 17:809643_G/A | 17 | 809643 | G | 0.0514 |
| 18q21.1 | rs11874392 | 18:46453156_A/T | 18 | 46453156 | A | 0.1606 |
| 19q13.43 | rs73068325 | 19:59079096_C/T | 19 | 59079096 | T | 0.0066 |
| 19p13.11 | rs34797592 | 19:16417198_C/T | 19 | 16417198 | T | 0.0824 |
| 19q13.11 | rs28840750 | 19:33519927_T/G | 19 | 33519927 | T | 0.1939 |
| 19q13.2 | rs1963413 | 19:41871573_G/A | 19 | 41871573 | A | 0.0441 |
| 19q13.33 | rs12979278 | 19:49218602_C/T | 19 | 49218602 | T | 0.0293 |
| 20q13.33 | rs2738783 | 20:62308612_T/G | 20 | 62308612 | T | 0.006 |
| 20q13.13 | rs6067417 | 20:48983697_C/T | 20 | 48983697 | C | 0.0331 |
| 20q13.12 | rs6031311 | 20:42666475_C/T | 20 | 42666475 | T | 0.0362 |
| 20q13.13 | rs6091189 | 20:49256285_C/T | 20 | 49256285 | T | 0.0549 |
| 20p12.3 | rs994308 | 20:6603622_C/T | 20 | 6603622 | C | 0.0626 |
| 20p12.3 | rs28488 | 20:6762221_C/T | 20 | 6762221 | T | 0.0714 |
| 20p12.3 | rs556532366 | 20:8568071_C/T | 20 | 8568071 | T | 0.0715 |
| 20p12.3 | rs189583 | 20:6376457_G/C | 20 | 6376457 | G | 0.0795 |
| 20p12.3 | rs4813802 | 20:6699595_T/G | 20 | 6699595 | G | 0.0819 |
| 20p12.3 | rs11087784 | 20:7740976_A/G | 20 | 7740976 | G | 0.0874 |
| 20q13.13 | rs6066825 | 20:47340117_A/G | 20 | 47340117 | A | 0.0719 |
| 20q13.13 | rs6063514 | 20:49055318_C/T | 20 | 49055318 | C | 0.0547 |
| 20q13.32 | rs13831 | 20:57475191_A/G | 20 | 57475191 | G | 0.0334 |
| 20q13.33 | rs1741640 | 20:60932414_T/C | 20 | 60932414 | C | 0.1146 |
| 20q11.22 | rs6058093 | 20:33213196_A/C | 20 | 33213196 | C | 0.045 |

Abbreviations: A, adenine; C, cytosine; G, guanine; T, thymine.

^a^ The missing reference single-nucleotide polymorphism was replaced by rs6904092 (linkage disequilibrium, D’=1 and r2=1).
