## Supplementary figures and images for "A single measurement of fecal hemoglobin concentration outperforms polygenic risk score in colorectal cancer risk assessment"

### Supplementary Figure 1

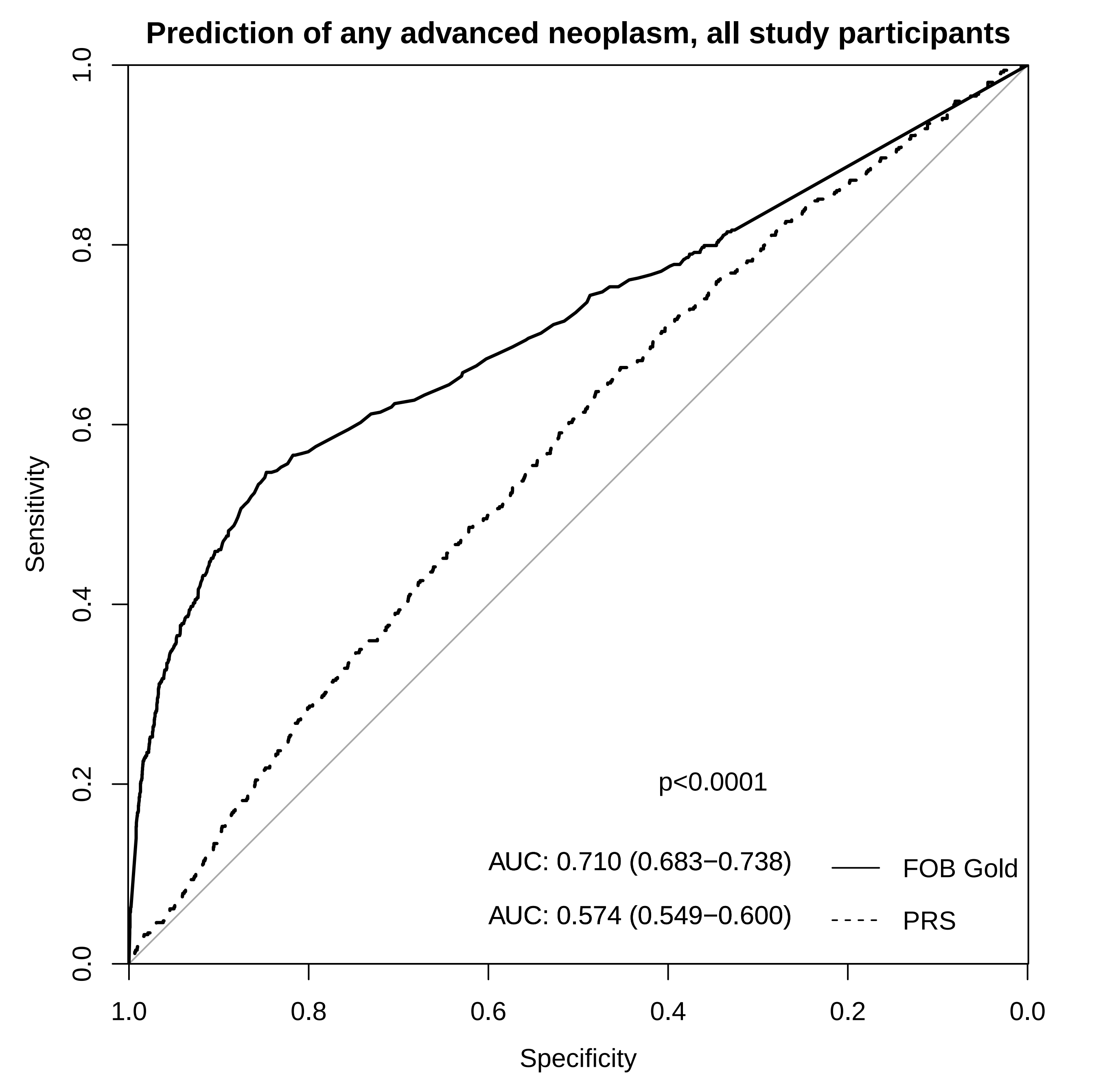
